# Factors influencing the commissioning and implementation of health and social care interventions for people with dementia: commissioner and stakeholder perspectives

**DOI:** 10.1101/2023.03.26.23287750

**Authors:** Rachael Tucker, Robert Vickers, Emma Adams, Clare Burgon, Juliette Lock, Sarah Goldberg, John Gladman, Tahir Masud, Elizabeth Orton, Stephen Timmons, Rowan H Harwood

## Abstract

**Background:** Despite several interventions demonstrating benefit to people living with dementia and their caregivers, few have been translated and implemented in routine clinical practice. There is limited evidence of the barriers and facilitators for commissioning and implementing health and social care interventions for people living with dementia. The aim of the current study was to explore the barriers and facilitators to commissioning and implementing a dementia friendly exercise and physical activity-based intervention (PrAISED (Promoting Activity, Stability and Independence in Early Dementia and Mild Cognitive Impairment)) in practice.

**Methods:** Qualitative semi-structured interviews were conducted with stakeholders from a range of backgrounds including individuals from universities, research centres, the voluntary and community sector, health and social care, and local government in England. The Consolidated Framework for Intervention Research (CFIR) was used to guide the design and analysis. Fourteen participants took part, including commissioning managers, service managers, partnership managers, charity representatives, commercial research specialists, academics/researchers, and healthcare professionals.

**Results:** Data were represented in 33 constructs across the five CFIR domains. Key barriers included cost/financing, the culture of commissioning, and available resources. Key facilitators included the adaptability of the intervention, cosmopolitanism/partnerships and connections, external policy and incentives, and the use of already existing (and untapped) workforces. Participants identified a need for greater support for people diagnosed with dementia and their caregivers immediately post dementia diagnosis.

**Conclusion:** Several barriers and facilitators for commissioning and implementing health and social care interventions for people with dementia were identified which need to be addressed. Recommended actions to facilitate the commissioning and implementation of dementia friendly services are: 1) map out local needs, 2) evidence the intervention including effectiveness and cost-effectiveness, 3) create/utilise networks with stakeholders, and 4) plan required resources.

## Introduction

Dementia is a neurodegenerative condition associated with a range of symptoms including memory loss, declining cognitive and executive function, and changes in behaviour and mood (Alzheimer’s Society, 2021). Over 55 million people are living with dementia worldwide (World Health Organisation [WHO], 2022) and this is projected to increase to approximately 152.8 million cases by 2050 (Nichols *et al.,* 2022). The global cost of dementia in 2019 amounted to US$ 1.3 trillion, with costs expected to exceed US$ 2.8 trillion by 2030 (WHO, 2022). Therefore, implementing interventions for dementia that focus on maintaining independence and slowing the rate of functional decline to prevent health and social care use and reduce this economic burden is important.

### Translating research into practice

The research into practice gap is well documented; it takes an average of 17 years for innovations to be implemented into routine clinical practice (Balas and Borden, 2000; Gitlin *et al*., 2020). Despite many non-pharmacological interventions for dementia demonstrating benefit, a small number are implemented in practice (Gitlin *et al*., 2020). Thus, it is crucial to understand strategies that facilitate their implementation.

There is little evidence for translating dementia friendly exercise/physical activity interventions into practice. A systematic review by Groot Kormelinck *et al*. (2021) identified barriers and facilitators for implementing complex interventions for residents with dementia living in long term care. In this review, only two interventions had an exercise or physical activity component (Groot Kormelinck *et al*., 2021), and due to its setting, may not have identified factors relevant to implementation across a range of health systems.

### Commissioning in England’s National Health Service

The National Health Service (NHS) in England is a publicly funded health system providing universal access to healthcare based on clinical need, not ability to pay (Department of Health and Social Care, 2021). NHS commissioning is complex whereby different services may be specified and paid for by different commissioners, including care systems, NHS England, Primary Care Networks and local government. Services may be provided by the voluntary and community sector (VCS), primary and secondary care health services, and support organisations working interdependently (Wenzel and Robertson, 2019; NHS England, 2021). However, social care in England is not universally funded, is commissioned by local government, provided by a range of providers, and is means tested (The Kings Fund, 2022). Commissioning dementia services is also therefore complex (National Institute for Health and Care Excellence [NICE], 2013). Such complexities mean many people with dementia and their families are often burdened with care costs and inadequate support (Alzheimer’s Society, 2018).

### The PrAISED Programme

The Promoting Activity, Independence and Stability in Early Dementia and Mild Cognitive Impairment (PrAISED) programme is a complex intervention which aims to keep people living with dementia independent and healthier for longer (Booth *et al*., 2018; Goldberg *et al*., 2019). PrAISED was developed by physiotherapists, occupational therapists, health psychologists, nurses, geriatricians, and carer representatives (Booth *et al*., 2018), and was tested in a feasibility study (Goldberg *et al*., 2019). It is a 12-month exercise and activity-based programme consisting of progressive strength training, balance exercises, functional activities and activities of daily living, dual task training, risk analysis, advice, and environmental assessment, all delivered using a motivational approach to support long-term participation in physical activity (Booth *et al*., 2018). The effectiveness of PrAISED was studied in the PrAISED-2 study was a multi-site, pragmatic, RCT, which took place between September 2018 and January 2023. The PrAISED-2 protocol can be accessed in full (Bajwa *et al*., 2019) and the results are forthcoming (Harwood *et al*.).

Given the lack of evidence for translation and implementation, and the complexities of commissioning, the current study aimed to identify barriers and facilitators for commissioning and implementing dementia friendly health and social care interventions in routine clinical practice using PrAISED as a case study, and to provide recommendations for future implementation.

## Methods

### Ethical approval

The study received research governance approvals and ethical approval from the Bradford Leeds Research Ethics Committee (18/YH/0059; 236099).

### Study Design

Qualitative, semi-structured interviews were used for data collection. A topic guide was developed which was informed by the Consolidated Framework for Implementation Research (CFIR) (Damschroder *et al*., 2009). Participants were asked to consider PrAISED in their answers, even if they had not been involved in the PrAISED RCT (see appendix one: interview topic guide).

### Participant Recruitment

Participants were stakeholders involved in the commissioning and delivery of dementia services. An introductory email was sent out to potential participants and/or contacts from pre-existing networks known to the research team. This included individuals working in the NHS/healthcare, social care, local authorities, the Voluntary and Community Sector (VCS), and other key stakeholder organisations concerned with commissioning, implementing, delivering, or promoting activity-based interventions for people with dementia and/or mild cognitive impairment; they were not necessarily involved in the PrAISED-2 RCT. Participants were provided with an information sheet and a consent form which was completed prior to their interview. An interview date and time was arranged to suit them. Two researchers (RT and RV) conducted the interviews, all of which were carried out, recorded, and transcribed using Microsoft Teams. Any identifiable information was removed from the transcripts and participants were assigned a participant number. A snowball sampling (or chain-referral sampling) technique was then employed to identify additional participants. These individuals were contacted by the research team via the introductory email and followed the same method of recruitment.

### Data Analysis

Data analysis was carried out using codebook thematic analysis (Braun *et al*., 2019). This type of thematic analysis uses a structured approach with predetermined themes and codes, or a research framework, to guide the analysis (Braun *et al*., 2019). This study used the CFIR (Damschroder *et al*., 2009) as a codebook. The CFIR was developed to consolidate published implementation theories into a consistent typology for use in evaluating implementation (Damschroder *et al*., 2009). Since its publication in 2009, the CFIR has grown in recognition and is now used widely across mixed method, quantitative and qualitative studies (Kirk *et al*., 2016). The CFIR consists of five domains: intervention characteristics, outer setting, inner setting, characteristics of individuals involved, and process of implementation (Damschroder *et al*., 2009). Across these domains are 39 constructs; full details are available at Damschroder *et al*. (2009) or https://cfirguide.org/.

Approximately halfway through the data analysis, revised CFIR guidelines were published updating constructs and their definitions (Damschroder *et al*., 2022). The methodological implications of this publication were considered collectively by the research team. After reviewing the updated framework, the team came to a consensus that due to time and funding constraints, the original CFIR would continue to be followed. As part of the updated CFIR, several constructs had their definition expanded or reworded (Damschroder *et al*., 2022). These new definitions were used in conjunction with the original CFIR codebook to guide coding decision-making, where the research team felt they better reflected uncoded data extracts. As per the updated CFIR framework, the research team worked collaboratively to define each domain in this study. The domains and their constructs used in this study are presented in Table 1. Constructs that were added or revised after the publication of the updated CFIR are denoted by *.

**Table 1.** CFIR domains and constructs (adapted from Damschroder *et al*., 2009 and Damschroder *et al*., 2022).

| Domain | Constructs |
| --- | --- |
| Intervention characteristics | Intervention source<br>Evidence strength and quality<br>Relative advantage<br>Adaptability<br>Triability<br>Complexity<br>Design quality and packaging<br>Cost |
| Outer setting | Patient needs and resources<br>Cosmopolitanism/partnerships and connections*<sup>N.B.</sup><br>Peer pressure<br>External policies and incentives<br>*Financing |
| Inner setting | Structural characteristics<br>Networks and communications |
|  | Culture<br>Implementation climate<br>- Tension for change<br>- Compatibility<br>- Relative priority<br>- Organisational incentives and rewards<br>- Goals and feedback<br>- Learning climate<br>Readiness for implementation<br>- Leadership engagement<br>- Available resources<br>- Access to information and knowledge |
| Characteristics of individuals | Knowledge and beliefs about the intervention<br>Self-efficacy<br>Individual stage of change<br>Individual identification with organisation<br>Other personal attributes |
| Process of implementation | Planning<br>Engaging<br>- Opinion leaders<br>- Formally appointed internal implementation leaders<br>- Champions<br>- External change agents<br>Executing<br>Reflecting and evaluating |
| N.B. Cosmopolitanism was renamed to reflect the nature of partnerships and connections as per the updated CFIR (Damschroder *et al.*, 2022). |  |

#### Data analysis process

NVivo software and a CFIR-approved pre-populated template (QSR International Pty Ltd. (2020) https://www.qsrinternational.com/nvivo-qualitative-data-analysis-software/home) were used to analyse the data; additional constructs were added where appropriate.

Braun and Clarke’s (2009) and Braun *et al*.’s (2019) thematic analysis steps were amended and/or combined to reflect the methods used in this study (codebook thematic analysis), which had predetermined codes and themes determined by the CFIR framework. Data analysis followed these steps:

1. Familiarisation (repeatedly reading transcripts and making notes about content)
2. Preliminary coding (preliminary coding into relevant constructs as per the CFIR codebook [available at https://cfirguide.org/tools/tools-and-templates/] and documenting rationale for coding decisions
3. Revising and revisiting coding/theme development (data revisited to check interpretations and amend if needed as researchers became more familiar with the data)
4. Finalising codes/themes (codes finalised within the research team)
5. Producing the report

Although these steps are presented as a sequence, data analysis followed an iterative process, with each step being revisited and revised. The lead author (RT) acted as lead coder for this study. A second coder (RV) reviewed a third of the transcripts to act as a peer-checker and reviewer of coding decisions. To improve understanding and collaborative use of the CFIR framework, the lead coder, second coder and wider implementation study team met weekly to discuss coding decisions.

## Results

A total of 14 participants took part in interviews. Participants included commissioning managers (n=4), service managers (n=3), charity representatives (n=1), partnership managers (n=1), commercial research specialists (n=1), academics/researchers (n=2), and healthcare professionals (n=2), working across a range of settings including universities, research centres, the VCS, health and social care, and local government. Interviews lasted between 25 and 68 minutes. Of the 40 constructs (39 original CFIR constructs, plus one from the updated CFIR (Damschroder *et al*., 2022) (Table 1), six had no entries during the analysis. These were: two constructs from the innovation characteristics domain (relative advantage and trialability), one from the inner setting domain (learning climate), two from the individual characteristics domain (self-efficacy and individual identification with organisation), and one from the process domain (executing). The remaining constructs were used as codes and were representative of extracts from the interview transcripts. The most frequently coded constructs were 1) needs and resources of those served by the organisation (outer setting), 2) available resources (inner setting), and 3) cosmopolitanism/partnership and connections (outer setting). Table 2 shows the frequency of coding for each construct (though frequency does not necessarily reflect importance), along with their classification as a barrier, facilitator, or both.

**Table 2:** Frequency of coding for each construct.

| Name | References | Barrier or facilitator |
| --- | --- | --- |
| **I. INNOVATION CHARACTERISTICS** |  |  |
| A. Innovation Source | 3 | Facilitator |
| B. Evidence Strength & Quality | 47 | Both |
| C. Relative Advantage | 0 | N/A |
| D. Adaptability | 21 | Facilitator |
| E. Trialability | 0 | N/A |
| F. Complexity | 1 | Barrier |
| G. Design Quality & Packaging | 2 | Facilitator |
| H. Cost | 42 | Barrier |
| **II. OUTER SETTING** |  |  |
| A. Needs & Resources of Those Served by the Organization | 187 | Both |
| B. Cosmopolitanism | 90 | Facilitator |
| C. Peer Pressure | 2 | Facilitator |
| D. External Policy & Incentives | 31 | Facilitator |
| Financing NB | 50 | Barrier |
| **III. INNER SETTING** |  |  |
| A. Structural Characteristics | 4 | Barrier |
| B. Networks & Communications | 9 | Facilitator |
| C. Culture | 16 | Barrier |
| D. Implementation Climate |  |  |
| 1. Tension for Change | 4 | Facilitator |
| 2. Compatibility | 6 | Facilitator |
| 3. Relative Priority | 11 | Both |
| 4. Organizational Incentives & Rewards | 12 | Facilitator |
| 5. Goals and Feedback | 2 | Facilitator |
| 6. Learning Climate | 0 | N/A |
| E. Readiness for Implementation |  |  |
| 1. Leadership Engagement | 3 | Facilitator |
| 2. Available Resources | 109 | Barrier |
| 3. Access to Knowledge and Information | 46 | Both |
| **IV. INDIVIDUAL CHARACTERISTICS** |  |  |
| A. Knowledge & Beliefs | 29 | Both |
| B. Self-efficacy | 0 | N/A |
| C. Individual Stage of Change | 2 | Facilitator |
| D. Individual Identification with Organization | 0 | N/A |
| E. Other Personal Attributes | 43 | Facilitator |
| V. PROCESS |  |  |
| A. Planning | 24 | Facilitator |
| B. Engaging |  |  |
| 1. Opinion Leaders | 3 | Facilitator |
| 2. Formally Appointed Implementation Leader | 1 | Facilitator |
| 3. Champion | 18 | Facilitator |
| 4. External Change Agent | 18 | Facilitator |
| 5. Key Stakeholders (Staff) | 23 | Facilitator |
| 6. Innovation Participants (Patients) | 3 | Facilitator |
| C. Executing | 0 | N/A |
| E. Reflecting & Evaluating | 9 | Facilitator |

As barriers and facilitators to the implementation of dementia friendly activity-based interventions were identified across all domains, this paper presents each domain and discusses barriers and facilitators within them, before presenting key meta-themes and considerations for the wider commissioning and implementation climate as part of the discussion.

### Innovation Characteristics

The innovation source, evidence strength and quality, adaptability, complexity, design quality and packaging, and cost, all represented barriers and facilitators. The PrAISED intervention was coproduced with patient and public representatives and healthcare professionals (Booth *et al*., 2018). Interviewees suggested coproduction was integral to successful implementation as the individual tailoring was seen to enhance participation, and the involvement of healthcare professionals provided reassurance of its effect:

> *‘…the fact that it’s also being developed with health professionals is something that’s really quite to its favour, because I think we find that people really look for reassurance from medical professionals, so if they know it’s got that medical endorsement, I think for us would be really positive,’* Participant 2 (Activity Manager).

Another facilitator was the innovation’s ability to be adapted to suit local systems. Several participants suggested that implementation would be facilitated and/or would be more likely to be commissioned if the innovation could be embedded within existing services:

> *‘I think if it’s something that you can almost add on to an existing provision … so you do have some of that skilled workforce, you have that management structure around it … some of the concerns of commissioners is when you end up with lots of small and then potentially vulnerable services … it just helps because you know you’ve got that capability there that could be mobilised rather than if you’re starting from scratch*,’ Participant 1 (Commissioner).

Some suggested utilising day services and/or care homes to deliver an intervention like PrAISED would keep costs down, utilise already existing services and upskill existing staff.

Another facilitator was the potential to use other professionals to deliver PrAISED in practice. In the main trial, PrAISED was delivered by occupational therapists, physiotherapists and rehabilitation support workers. Participants in the current study suggested other professionals, such as exercise instructors, could take on responsibility for delivering a dementia friendly, exercise-based intervention and would be qualified to do so (discussed in greater depth in the individual characteristics domain). Interestingly, this view differed from those of healthcare professionals interviewed as part of a pilot PrAISED service, who felt it was essential healthcare professionals delivered exercise interventions for people living with dementia (Adams *et al*., forthcoming). It was suggested this potential adaptation had collateral benefits for cost, and could reduce the demand on the existing workforce, utilise an untapped workforce and improve collaborative working with the local community, for example, leisure centres.

Evidence was a significant factor and facilitator in the commissioning of an intervention like PrAISED:

> *‘It’s an area that you’ve got to have as much efficacy, evidence as possible… that is what is going to determine the success,’* Participant 7 (Commercial Director).

> *‘If the evidence isn’t there to support it, then it’s not going to be there ultimately,’* Participant 12 (Partnership Manager).

One strand of evidence that was particularly pertinent to successful commissioning was the intervention’s ability to deliver cost savings and where these would be visible, for example, health versus social care. However, this was deemed difficult to evidence. Participants 13 and 14, both commissioning managers, described the importance of interventions delivering cost savings in influencing decisions and allocating funds:

> *‘If we can start to evidence that this is delaying or improving outcomes… I think that would help massively… It’s like that invest to save sort of thing, isn’t it? If we can really show some evidence around that… what support around this does, then I think that you’ve got more of a chance,’* Participant 13 (Commissioning Manager).

> *‘Delaying need for social care is a really big thing for us. So, if an organisation came and said look, we can prevent people hitting your services for a long time, that’s a really big driver for us, and like promoting independence, so, even if people are using our services, they’re using them less and living at home longer,’* Participant 14 (Commissioning Manager).

Also, participants working outside of commissioning recognised how crucial evidence was to the decision-making process:

> *‘It’s also important to show that there’s evidence, which obviously PrAISED is there to do… certainly some people in commissioning are a bit swayed by evidence or are very sceptical about things unless there’s evidence,’* Participant 8 (Professor of Dementia Research).

Interestingly, participant 7 described how different ‘levels’ of evidence would be required, depending on the system of delivery. For example, lower-level evidence would be required if the intervention were to be self-funded, as it would be an *‘emotional purchase’* by family members and/or carers, whereas:

> *‘…if it’s a statutory service provision model [local authority or NHS], then the bar is higher in terms of the amount of certainty that they would need in order to commission it and that might be certainty around patient outcomes, deferred benefit, cost versus benefit, cost benefit analysis. They’re going to want to see that and understand it because with limited budgets and competing demands for resources, they want to put their bets on the horses that are going to get them the biggest returns. Otherwise, it might fall into that nice to have, but not essential, which is really hard,’* Participant 7 (Commercial Director).

Whilst evidence of RCT outcomes was mostly advocated, other forms of evidence, such as qualitative research, were also important:

> *‘… it’s about showing real life stories and the positive impact it can have on someone’s life… it’s actually sharing real life people and how it has impacted on their life and what they’ve been able to go on to do after they’ve had that intervention. I think that’s really powerful,’* Participant 6 (Sports Development Officer).

### Outer setting

Most participants reported that there is a need for dementia friendly activity-based interventions. Participants recognised the benefits of physical activity, and many proactively promoted this. Some reported that there were vast amounts of initiatives which aimed to engage people with long term conditions in physical activity and exercise. However, importantly, these were mostly deemed unsuitable for people living with dementia:

> *‘You need to have sort of a specific understanding of their needs and what’s going to be most likely to support them into activity and help them to maintain that… often people with dementia, when we’re talking to them about some of the services and support that we’re providing, they find it a little bit harder to relate to some of the messaging and a bit harder to undertake some of the activities… they need to be communicated in a particular way and they need to take into consideration their ability level and just them as a whole person,’* Participant 2 (Activity Manager).

There were few dementia specific or dementia friendly services currently being provided, though participant 10 reported that there was ‘*an appetite definitely to improve the provision or enhance the provision or create the provision to start with*.’ Participants described the post-diagnostic support as lacking, and at worst, absent:

> *‘We have a gap… the post-diagnostic offer to people with dementia is pretty woeful,’* Participant 8 (Professor of Dementia Research).

Participants described efforts in their organisation and/or local area to provide or promote dementia friendly interventions, such as dementia friendly swimming and golf. However, what was evident across the data was an identified need to map what was already available, and to evaluate the needs of the local population living with dementia, including marginalised and underserved communities. Participants attempted to address unmet need and deficits in specialist dementia knowledge through training and education for care home and day centre staff, and dementia specialist accreditation. Some described using roles such as social prescribing (referrals from healthcare professionals to local non-clinical services [e.g., volunteering, sports groups etc] with the aim of holistically improving health and wellbeing [Buck and Ewbank, 2020]) to engage this population in exercise, and others created dementia hubs and strategies to support local priorities. Participants identified several barriers to engaging their local community of people living with dementia in physical activity. This included fear and anxiety, avoidance of perceivably risky activity, lacking support, poor awareness of available services, and lacking infrastructure and transport links, which were troublesome in rural areas.

Participants considered an intervention like PrAISED to be an important component in addressing the post-diagnostic support gap which could play an important role in preventing health and social care use. This was a particular concern in the face of exponential growth in the number of people living with dementia. For some, this underpinned the demand for services like PrAISED:

> *‘I think it is critical because we are very limited in the resources we have, so everything you can do to keep people at the lower levels of care for as long as possible are critical and keep people in their own homes wherever possible. So yeah, absolutely. I think anything that supports that kind of left shift to our demand management is really critical,’* Participant 1 (Commissioner).

Early support was deemed necessary to not only prevent health and social care consumption, but also to enhance quality of life and promote meaningful activity and engagement in all aspects of life.

A significant facilitator to providing dementia friendly services was collaborative working and the formation of partnerships and connections with other organisations and stakeholders. Participants were hopeful the recent change from Clinical Commissioning Groups (CCGs) to Integrated Care Systems (ICS) would improve collaborative working and align commissioning priorities across health and social care in England. CCGs were responsible for planning and commissioning health services in their local area and in July 2022, they were abolished and replaced by ICSs, consisting of partnership organisations working collaboratively to join up health and care provision in their local area (NHS England, 2022). Despite optimism regarding these new partnerships, there was confusion surrounding the responsibilities of these groups and concerns that this would complicate the commissioning process. Additionally, competing priorities between organisations attempting to work cohesively posed a challenge.

Nonetheless, these partnerships were imperative to effective commissioning. Most participants emphasised the importance of the voluntary sector in the provision of dementia friendly services (if commissioned to do so). Many stakeholders had experience working with charities in the design, delivery and maintenance of dementia services and they advocated for their presence as specialists in dementia. Some suggested these organisations were best placed to deliver services (if commissioned) as they had the time, resources, and specialist knowledge to do so. Alongside charities and the voluntary sector, stakeholders described collaborations with national sporting agencies such as Sport England and other partnerships, including universities, place-based partnerships, social enterprises, the Fire and Rescue Service, community groups, commercial advisors, professional sports teams, and health and social care organisations. These partnerships were seen to facilitate service sustainability and long-term presence in the community.

Organisational partnerships also facilitated the financing of dementia friendly services. These organisations had grants which could fund services, though these were often short lived. Financing was a significant barrier to the commissioning and implementation of dementia friendly interventions. There were tensions between the responsibility for funding:

> *‘Personally, I think [the] NHS should give us money towards it if they want us to implement it… it will have a knock-on effect on the admissions because if we reduce falls for a longer period of time, it means they’ve got less operations to do and less throughput of hospitals,’* Participant 4 (Occupational Therapist).

The private versus public funding debate was influenced by several factors. Some reported private financing of services was a feasible method for delivering interventions like PrAISED. In contrast, public funding was regarded as difficult to obtain and was frequently linked to other constructs, such as external policy and incentives, and available resources in the inner setting. The VCS thus frequently bridged the gap, and there was a reliance on this sector, which was not without consequences:

> *‘It is a difficult one because it it’s one of the areas where there is a lot of reliance on almost free services as in non-funded services so that they’re either a charitable or community, so church or other kind of charity type organisations providing things, which means it’s quite piecemeal and quite localized. So, it’s quite hard,’* Participant 7 (Commercial Director).

In terms of what drove the commissioning and implementation of dementia friendly services in the outer setting, there was little reference to peer pressure, though participant 9 highlighted the importance of being aware of what competing organisations were doing and what services were already available. A more commonly cited construct was external policy and incentives. There were conflicting views on the value of external policy and incentives in influencing the commissioning and implementation of dementia friendly services, where it was seen as sometimes a facilitator and at other times, non-influential:

> *‘We always say “oh policy drives action,” but it doesn’t always… if I was building a business around this, I wouldn’t be relying on policy to be the driver… At the end of the day, policy is slightly important… this is my own view, [NHS] Trusts tend not to buy things because of policy. Trusts buy things because it solves a problem for them,’* Participant 7 (Commercial Director).

However, other participants felt policy acted as a facilitator:

> *‘The easiest way to get it funded is where actual national policy says you must have X service in place. That’s the easiest thing. And you have ring fenced money. I think it’s really hard if you don’t have that… if we’ve got a national policy, we do have to respond to it,’* Participant 1 (Commissioner).

They went onto suggest external monitoring, performance management and Key Performance Indicators (KPIs) also facilitated commissioning.

Local strategy and policy were also seen to both facilitate and hinder implementation, as budgets would be allocated accordingly:

> *‘I think probably the one of the main factors is it being a strategic priority locally, because then you’ve got the buy in from the whole system and at the top. So, if it ain’t a strategic priority, then even if it is really good, it might not continue to be funded because of the things which are meeting those strategic priorities will likely get more resources allocated because budgets will be allocated on what are those strategic priorities,’ Participant 5* (Commissioning Manager).

Financial incentives and penalties which are used across OECD (Organisation for Economic Co-operation and Development) member countries to motivate performance in health systems (Milstein and Schreyoegg, 2016) were also perceived facilitators:

> *‘I suppose targets and financial incentives or financial pen-well incentives are better than penalties, but usually in the NHS is about punishment. So, you know some sort of stimulus that’s hard for them to ignore. So simply giving them advice that they should is “well, we can ignore that then.” So, it needs to be a bit of force behind it to make people actually implement things,’* Participant 8 (Professor of Dementia Research).

### Inner setting

Participants described a need to shift the culture of commissioning from short to long term. Several participants expressed concern that commissioners focussed on ‘crisis management’ due to the NHS climate, rather than on preventative interventions that would provide cost efficiency savings longer term. It was perceived as more difficult to achieve buy-in to such interventions, as often cost savings were not immediately visible. Physical activity and public health interventions were perceived as key to preventative care, and whilst there was a shift towards these types of interventions, there was still work to be done:

> *‘…in terms of how much we value we place on physical activity in terms of prevention and treatment for long term conditions… I don’t think we’re quite where we should be with that… the health service has been increasingly crisis weighted and I think that limits how much we think about building in preventative or wellbeing factors into primary services,’* Participant 12 (Partnership Manager).

There was a shift in culture towards collaborative working, both within the inner setting (networks and communications), and outer setting (cosmopolitanism/partnerships and connections). However, inner setting decision making processes remained complex and at times, posed a barrier to commissioning and implementation. Indeed, for participant 11, they had observed how networks facilitated implementation, but also introduced biases, causing them to question the system:

> *‘I seem to find if they like something and they have a good relationship with an organisation, funnily enough, that sometimes leads to funding and renewal of funding… it would be nice to think it is a fair process that looks at the pros and cons and what’s best for the individual, but I think with a lot of things particularly that are NHS system based is that they’re very rigid in what they want them to achieve and although they may say that they’re person-centred, really, they’re system-centred and then the person is expected to fit in with that,’* Participant 11 (Researcher).

As described earlier, there was an identified need for dementia friendly activity-based interventions. For three participants, their views met the criteria for coding under the construct tension for change, as they viewed the current situation as intolerable or requiring urgent change. Nonetheless, this was subject to challenges. It was important for any innovation attempting commissioning and/or implementation to be compatible with the existing local systems. For example, whether the innovation could be embedded or absorbed into existing services (compatibility), which is linked to the adaptability construct (innovation characteristics). This was a significant facilitator for implementation success.

Furthermore, the relative priority of the innovation was both a barrier and facilitator. Priorities within the commissioning cycle could prevent similar services from being commissioned. For example, participant 5 suggested that if a falls prevention programme had recently been commissioned, other dementia friendly activity-based interventions would be a lower priority for commissioning. Moreover, the wider social, political, and economic climate also shifted commissioning priorities; the most recent example being the COVID-19 pandemic, where public health and pandemic management were inevitably given greater priority. Furthermore, organisational rewards, measurement and KPIs acted as incentives to implement innovations, but only if local priorities and strategies deemed dementia care and falls prevention a priority. More so, should the innovation align with the goals and mission statement of the organisation, this too would escalate the priority of commissioning and implementation.

One of the most significant and highly cited barriers to commissioning and implementing dementia friendly services was a lack of available resources. This included workforce, time, capacity, available providers, and most significantly, funding. Appropriate (and long-term) funding to commission, implement and deliver an innovation was difficult to secure. Often, budgets were already allocated and thus, unavailable:

> *‘The real challenge we have got of course is there isn’t new money, there isn’t spare money,’* Participant 1 (Commissioner).

Considering the vast array of contextual factors represented across the CFIR constructs, it is significant that participants often came back to the topic of resources. This issue was shared across the stakeholders, including those with commissioning responsibilities, who expressed frustration that they were unable to commission innovations:

> *‘There isn’t a lot of money… this is a really frustrating thing that you get all these people coming to you with some really good things [innovations], but we don’t really have money for spending on these things anymore,’* Participant 14.

In the context of limited resources, the NHS was suggested to be the most suitable provider of a service like PrAISED:

> *‘The problem for dementia is that much of it falls between health and social care. Social care is so poorly funded that it is difficult to see it doing a great deal… probably for it to become more widespread the way things currently are, it would require NHS commissioning, I think are the only people with any money,’* Participant 8 (Professor of Dementia Research).

In addition to funding, inadequate staffing levels and capacity of existing staff hampered implementation. Staff would be required to take on additional workload or redirect time from other services to implement innovations, which was undesirable. This was also the case for allocating time for training. Some suggested additional staff could be hired to facilitate implementation; however, this was associated with greater costs, temporary contracts, and thus, job insecurity. The demands of a lengthy programme like PrAISED (delivered over 12 months) was deemed unfeasible, as participant 4 described when looking to implement Otago, a home-based balance and strengthening programme effective at reducing falls in over 65s (Campbell and Robertson, 2007):

> *‘The main thing is time and follow ups. We just can’t… Otago’s 12 months. We can’t do it. We can’t do it,’* Participant 4 (Occupational Therapist).

Although leadership engagement (such as service managers) could facilitate this, resources frequently dictated the success of commissioning and implementing innovations.

### Individual characteristics

The characteristics of individuals responsible for commissioning, implementing and delivering interventions like PrAISED, acted as potential facilitators to success. Participants identified areas where knowledge could be instilled to upskill caregivers (formal and informal) to engage people living with dementia in physical activity interventions. Furthermore, the knowledge of and belief in such interventions acted as a driver. Individuals’ stage of change (Damschroder *et al.,* 2022) thus could initiate service development; for example, when asked what a persuading factor in the commissioning and implementation of a dementia friendly intervention could be, participant 6 stated:

> *‘I wouldn’t need persuading because I’m completely on board with it,’* Participant 6 (Sports Development Officer).

As mentioned earlier (innovation characteristics), many participants suggested the intervention could be delivered by other professional groups, such as exercise instructors, personal trainers, domiciliary care workers and support workers/therapy assistants. This was captured under the other personal attributes construct of the individual characteristics domain. Professional groups outside of physiotherapy and occupational therapy were suggested as potential deliverers of interventions like PrAISED due to their cheaper cost, connections to local communities (e.g., gyms, leisure centres, community groups), and their perceived undervalue as an untapped workforce. Furthermore, difficulty in recruiting clinicians and the pressure existing clinicians were under was acknowledged and thus, alternative groups taking responsibility for an intervention like PrAISED would ease pressure.

Most participants expressed a growing appreciation of exercise professionals in delivering physical activity interventions:

> *‘…there are thousands of physical activity exercise professionals who are highly qualified… Let’s use that workforce. Why not? You know, they are an untapped workforce and there’s a lot of them out there who are already got those connections in the community… they’ve got those behaviours, skills and those motivational interviewing techniques to work with those individuals and then perhaps to support the carers directly as well as those are being cared for. So huge opportunities there,’* Participant 10 (Project Manager).

Many suggested these members of the workforce were qualified and competent to deliver an intervention like PrAISED, with many having undergone specialist training in long term conditions. Thus, it was not always deemed necessary to have registered clinical qualified healthcare professionals’ oversight, though some suggested clinicians could work collaboratively to oversee the programme with exercise professionals delivering the intervention. The use of an existing, untapped workforce could impact the success of commissioning, though this had implications for the intervention:

> *‘…with all the pressures in the system, with workforce, the interventions that can be delivered successfully, carefully, safely, but with the lowest level of staff training required are very appealing… depending on the intervention that depends how achievable that might be, but that’s where I would be looking at… what is the lowest level of staff that you could utilize on this without making it unsafe or ineffective?’* Participant 7 (Commercial Director).

Some suggested having non-registered clinical staff delivering the intervention would be the most realistic option for commissioning and implementing a service such as PrAISED.

### Process

References to planning the implementation process were mostly dominated by the planning of commissioning. As this work package was not reflecting retrospectively on an implemented service, participants spoke hypothetically about this process. The greatest concern was how to plan the business case or model to facilitate successful commissioning/securement of funding. These concerns were mostly related to other constructs such as financing (outer setting) and available resources (inner setting). Other concerns were regarding the organisational model within local systems, such as commercialisation and licencing and how these would be managed in the future, as this had implications for an intervention’s sustainability. Additionally, participants suggested it was imperative to be cognisant of the commissioning cycle and plan attempted business cases accordingly, as this could affect success. Participant 5 described it as being *‘in the right place, at the right time.’*

In the case of the English NHS, having a range of engaged individuals was integral to implementation success. Participants provided several examples, including opinion leaders (e.g., leaders in dementia research, dementia advocates), formally appointed implementation leaders (e.g., project leads, healthy aging leads), external change agents (e.g., opinion leaders, politicians, councillors, commissioners, advisors, television personalities^i^), champions (self and/or formally appointed), key stakeholders (healthcare professionals, staff, organisations), and innovation participants (service users and caregivers). These champions were considered key to driving the implementation process, particularly when faced with challenges or decreasing momentum:

> *‘We do need to have if you want to call [them] falls, champions or dementia champions, if that’s the right word, but more ambassadors or business change agents…. Within those day services who can take a bit of ownership and accountability to ramp up that effort,’* Participant 9 (Programme Manager [Commissioning]).

> *‘…it’s enthusiasm and passion for me that’s such an important driver,’* Participant 11 (Health and Activity Researcher).

Reflection and evaluation were critical parts of the implementation process for some participants and was something that needed to be built in as part of the planning process. This was important to not only evaluate implementation success and ‘continuous improvement,’ but to provide lessons for future implementation.

## Discussion

### Summary

The aim of the current study was to explore the barriers and facilitators to commissioning and implementing a dementia friendly exercise and physical activity-based intervention in the English health and social care system using PrAISED as a case study. We found facilitators and barriers mapped onto the CIFR which showed:

1. The credibility and cost-saving nature of the intervention was important, along with the ability to adapt it to local provision and skill mix.
2. Interventions such as PrAISED may fill the post diagnostic gap, but there needs to be an organisational system that will get them commissioned; this involves collaboration between commissioners, providers and other stakeholders.
3. There also needs to be a policy culture that values prevention, prioritises dementia and is willing to commit resource to it to make it work.

### The post diagnostic gap

The post-diagnostic gap is defined as *‘an umbrella term encompassing the variety of official and informal services and information aimed at promoting the health, social, and psychological wellbeing of people with dementia and their carers after a diagnosis. Integrated treatment, care, and support are the pillars of effective post-diagnosis models,’* (World Alzheimer Report, 2022, p.21). This was a common theme in this study and is a global problem (World Alzheimer Report, 2021; Gresham, 2022), despite efforts designed to address this (Barrett and Burns, 2014; NICE, 2018).

Consequently, there is a need for innovations that address the service gap. Many participants advocated for physical activity interventions, though they also identified a broader need for psychosocial, emotional, logistical, practical, and peer support. This echoes the findings of Bamford *et al*. (2021), who identified 20 components of post diagnostic support, extending across five themes (timely identification and management of needs; understanding and managing dementia; emotional and psychological wellbeing; practical support; and integrating support). Bamford *et al*. (2021) suggested there is a need for local planning and coordination of such services, and there was evidence of this in this study, though wider barriers to commissioning and implementation had the potential to hamper efforts.

This study’s findings reflect other literature exploring barriers and facilitators to commissioning and implementing post-diagnostic services. Wheatley *et al*. (2021) identified unsupportive infrastructure, limited proactive, holistic tailored support, and limited capacity and capability as barriers to implementation. They identified strategies to address this, such as creating opportunities for service improvement, facilitating collaborative working, supporting non-specialists (e.g., non-medically qualified healthcare professionals) to deliver dementia care, and the development of ongoing holistic support (Wheatley *et al.,* 2021). The current study provides evidence that these strategies are being undertaken, though there is more to be done to enhance collaboration and the utilisation of existing workforces.

Some research suggests that physical activity interventions for older people can be delivered safely and effectively by non-clinically registered professionals (e.g., exercise instructors, postural stability instructors) (Iliffe *et al*., 2014) and can be delivered in novel environments outside of traditional healthcare settings (Long *et al*., 2020). Furthermore, a physical activity intervention for older people with cognitive impairment, delivered by exercise instructors, showed promising improvements in physical and cognitive function, quality of life and caregiver burden, though the sample size was small (Barnes *et al*., 2015). Therefore, the delivery of physical activity interventions by these professionals may offer a solution to the commonly cited barrier of available resources, which was recommended by Wheatley *et al*. (2021).

### The culture of commissioning in England

This study identified the need for a policy culture that values prevention. In the UK, prevention of ill health is described as a role for individuals, communities, NHS, social care, and local and national government (Department of Health and Social Care, 2018), and is a global priority (WHO, 2019). However, these findings demonstrate the complexities of prevention in practice in a universal publicly funded health system.

Interventions like PrAISED are preventative and they were considered harder to commission. Participants suggested this was twofold: 1) the benefit of such interventions was not immediately visible, and 2) commissioning was focussed on short term ‘crisis management.’ Participants suggested the underappreciation of preventative services meant interventions that may provide longer term cost savings were harder to gain support for and thus implement. This was coupled with difficulty in evidencing cost savings, particularly as commissioners wanted to be able to evidence specifically where cost savings would be delivered, e.g., health or social care. Despite this, participants with commissioning responsibilities were generally acutely aware of the need for preventative services, with some creating dementia strategies and influencing local priorities to address this. Nonetheless, this has the potential to create fragmentation and inequity across sectors and geographies. Furthermore, despite actions to address this, commissioners were also subject to the barriers to commissioning and implementation identified in this study.

In the wider literature, the discourse surrounding joint commissioning emphasises prevention (Dickinson *et al*., 2013). Miller *et al*. (2013) suggest delaying deterioration and maintaining physical and mental health in older people (and thus, their use of health services) is a commonly cited aspiration in commissioning (e.g., Allen and Glasby, 2010). However, it appears the ability to exercise this rhetoric is limited in the face of competing priorities and restricted resources (affordability). In this study, the VCS was seen as an able facilitator and provider of preventative care, something earlier suggested by Miller *et al*. (2013). While there have been successful examples of this, the issues with demonstrating preventative and rehabilitative services, as well as the need to rebalance the system with such care being integrated (Allen and Glasby, 2010), continue to pose challenges in commissioning.

The current study has considered the commissioning and implementation of dementia friendly exercise and physical activity-based interventions using PrAISED as a case study. It has identified key considerations for the future of dementia care, particularly in relation to provision of post diagnostic support and the culture of commissioning in contemporary healthcare. Furthermore, it has identified barriers (cost/financing, the culture of commissioning, and available resources) and facilitators (adaptability of the intervention, cosmopolitanism/partnerships and connections, external policy and incentives, and use of already existing workforces) to commissioning dementia friendly services. Thus, this study provides insight for stakeholders planning the commissioning, implementation and promotion of dementia services.

### Recommendations for commissioning and implementing dementia services

A series of recommendations have been collated based upon the barriers and facilitators identified in this study:

1. Map out local needs and resources
  a. The needs and resources of the population living with dementia and their caregivers should be identified (including the needs of underserved communities)
    i. Involve people living with dementia and their caregivers in identifying these needs
  b. Map existing services (and how/where the intervention would fit)
2. Evidence the intervention
  a. Evidence the outcomes of the intervention, including effectiveness and cost-effectiveness (e.g., physical and mental health, psychosocial factors, and financial such as cost benefit analysis, patient and deliverer satisfaction [e.g., qualitative data]), to ensure stakeholders value the innovation and its potential impact to ensure it is commissioned/funded and integrated into routine clinical practice.
3. Create/utilise networks and partnerships with stakeholders with a role in implementing, commissioning, providing, and promoting dementia friendly interventions
  a. Identify local/organisational priorities, resources, and opportunities for collaboration to facilitate commissioning and implementation
  b. Involve these networks and partnerships in the early stages to plan for sustainability
4. Plan required resources for delivery (cost, staffing, equipment)
  a. Assess capacity in the local system for non-medical professionals delivering exercise and physical activity interventions (e.g., exercise instructors), where able to do so safely and appropriately.

### Strengths and limitations

This study presents the perspectives of a small number of stakeholders thus they will not necessarily represent the views of all stakeholders involved in dementia care or commissioning. As this study was carried out in England, the views may not be representative of stakeholders in other countries and care systems.

A strength of the study was the range of perspectives and expertise collected, as all participants were involved in dementia services commissioning and provision.

Furthermore, the collective discussion of coding decisions within the wider implementation research team meant a range of perspectives were utilised during data analysis.

## Conclusion

This study identified several barriers and facilitators to the commissioning and implementation of dementia friendly exercise and physical activity-based interventions such as PrAISED. Key barriers to commissioning and implementing dementia specific services included their cost/financing, competing commissioning priorities and having available resources. Key facilitators included the adaptability of the intervention, having good partnerships and connections in place, external policy and incentives, and the use of already existing (and untapped) workforces.

Based on the results of this study, four actions are recommended to facilitate the commissioning and implementation of interventions like PrAISED: 1) map out local needs and resources, 2) evidence the intervention including effectiveness and cost-effectiveness, 3) create/utilise networks with stakeholders, and 4) plan required resources. Further research is required to explore the outcomes of proposed recommendations.

## Data Availability

All data produced in the present study are available upon reasonable request to the authors.

**Appendix One:**
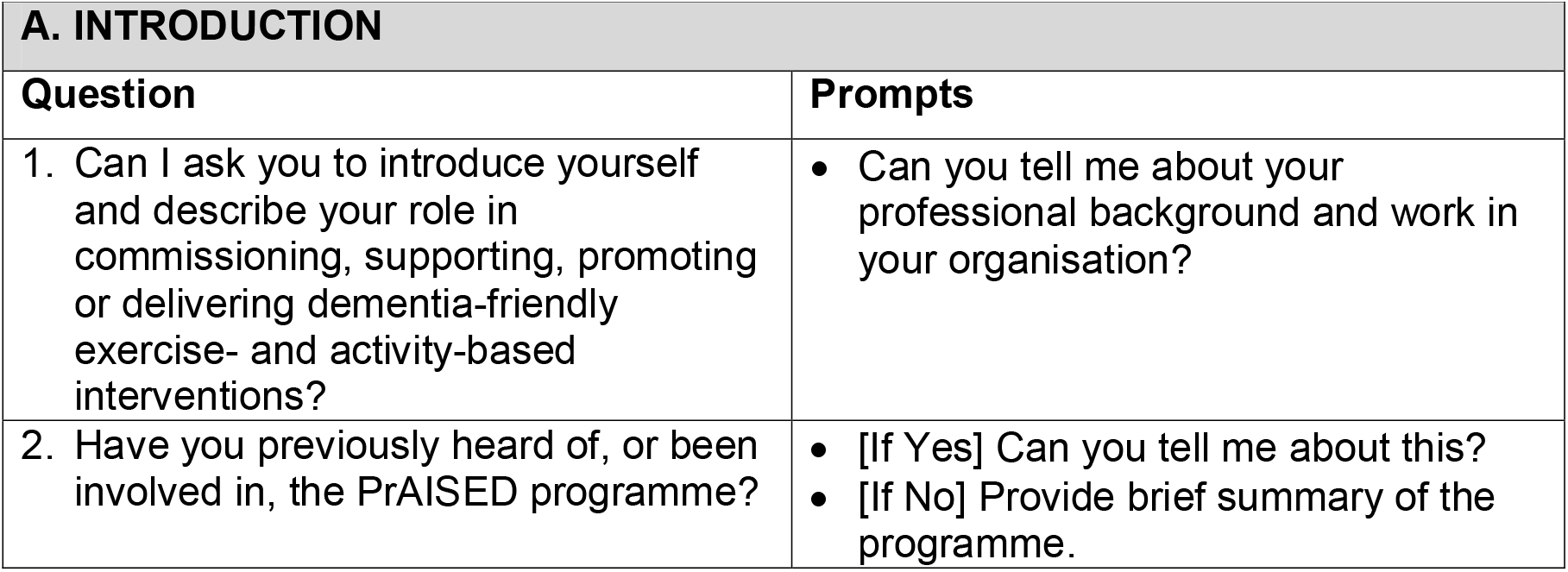
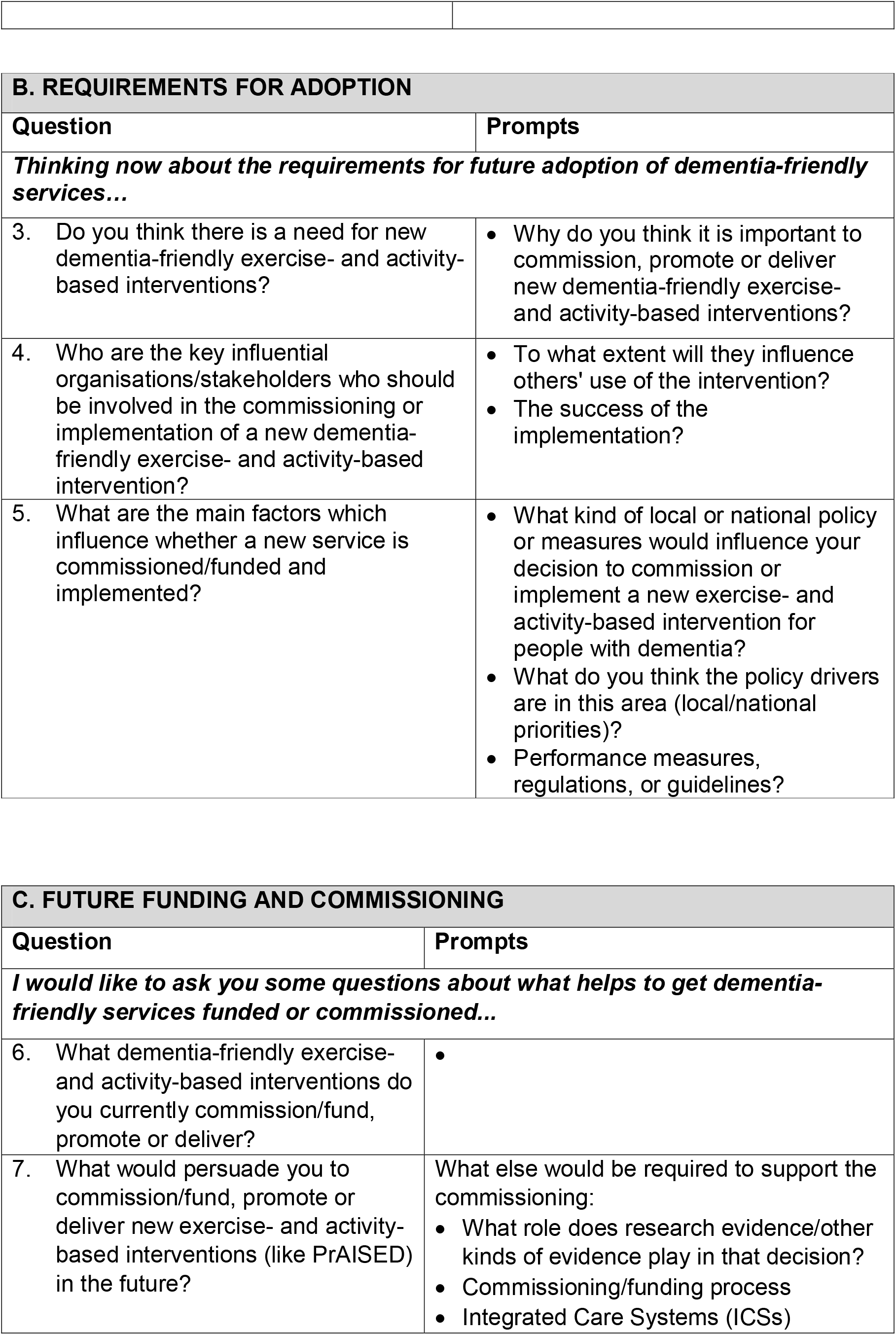
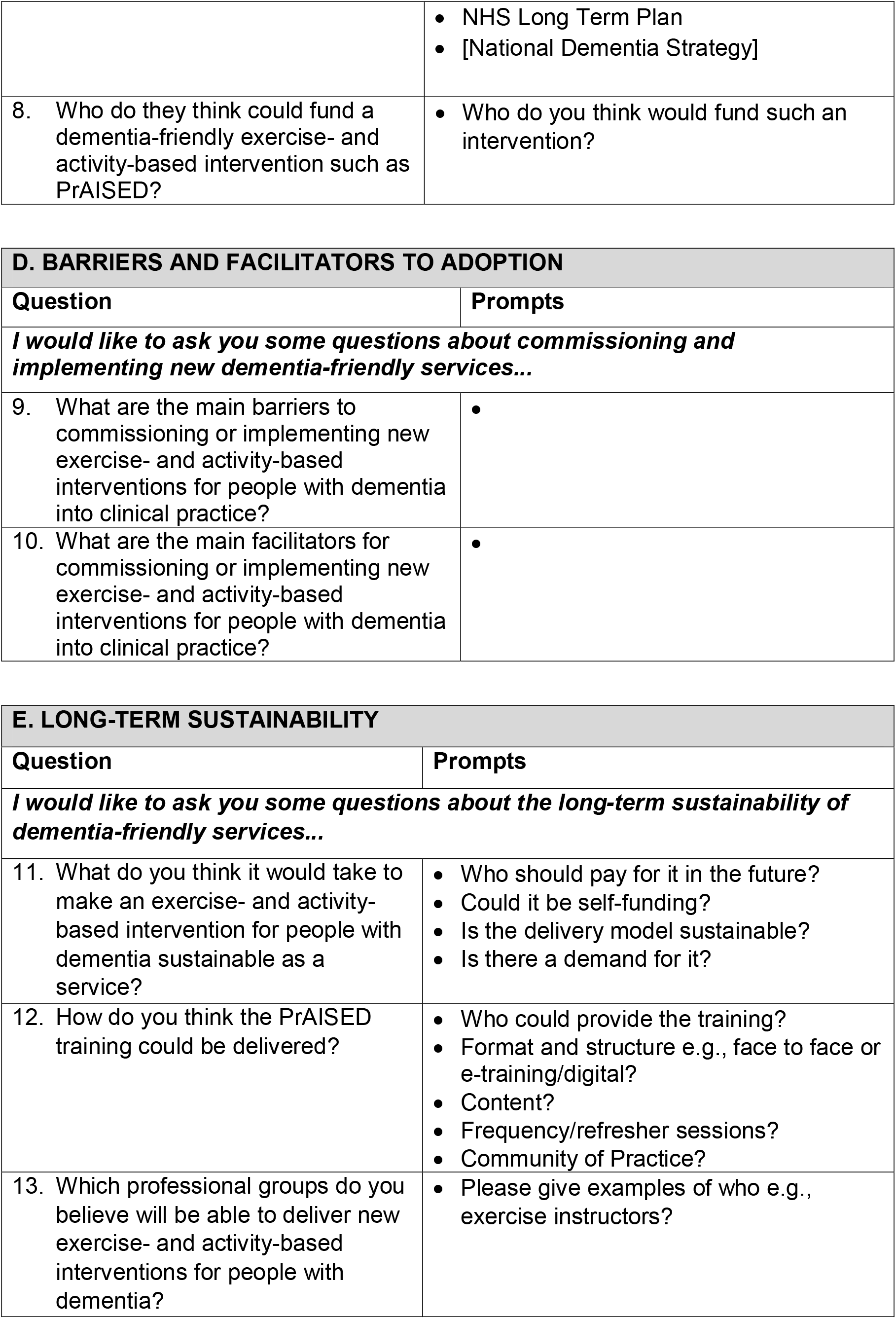
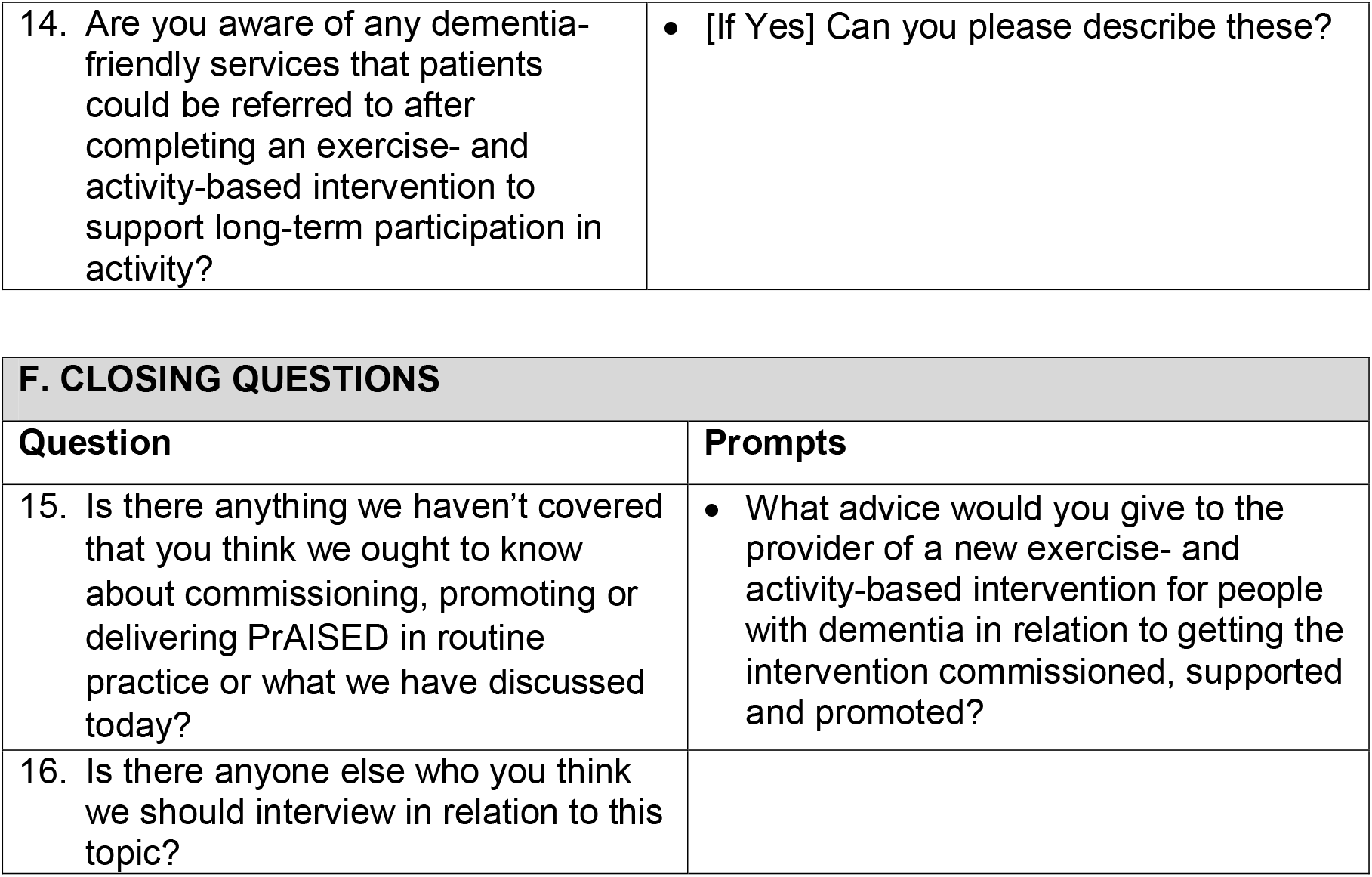
Interview Topic Guide.

| A. INTRODUCTION |  |
| --- | --- |
| Question | Prompts |
| 1. Can I ask you to introduce yourself and describe your role in commissioning, supporting, promoting or delivering dementia-friendly exercise- and activity-based interventions? | • Can you tell me about your professional background and work in your organisation? |
| 2. Have you previously heard of, or been involved in, the PrAISED programme? | • [If Yes] Can you tell me about this?<br>• [If No] Provide brief summary of the programme. |

| **B. REQUIREMENTS FOR ADOPTION** |  |
| --- | --- |
| **Question** | **Prompts** |
| ***Thinking now about the requirements for future adoption of dementia-friendly services...*** |  |
| 3. Do you think there is a need for new dementia-friendly exercise- and activity-based interventions? | • Why do you think it is important to commission, promote or deliver new dementia-friendly exercise- and activity-based interventions? |
| 4. Who are the key influential organisations/stakeholders who should be involved in the commissioning or implementation of a new dementia-friendly exercise- and activity-based intervention? | • To what extent will they influence others' use of the intervention?<br>• The success of the implementation? |
| 5. What are the main factors which influence whether a new service is commissioned/funded and implemented? | • What kind of local or national policy or measures would influence your decision to commission or implement a new exercise- and activity-based intervention for people with dementia?<br>• What do you think the policy drivers are in this area (local/national priorities)?<br>• Performance measures, regulations, or guidelines? |

| **C. FUTURE FUNDING AND COMMISSIONING** |  |
| --- | --- |
| **Question** | **Prompts** |
| ***I would like to ask you some questions about what helps to get dementia-friendly services funded or commissioned...*** |  |
| 6. What dementia-friendly exercise- and activity-based interventions do you currently commission/fund, promote or deliver? | • |
| 7. What would persuade you to commission/fund, promote or deliver new exercise- and activity-based interventions (like PrAISED) in the future? | What else would be required to support the commissioning:<br>• What role does research evidence/other kinds of evidence play in that decision?<br>• Commissioning/funding process<br>• Integrated Care Systems (ICSs) |
|  | • NHS Long Term Plan<br>• [National Dementia Strategy] |
| 8. Who do they think could fund a dementia-friendly exercise- and activity-based intervention such as PrAISED? | • Who do you think would fund such an intervention? |

| **D. BARRIERS AND FACILITATORS TO ADOPTION** |  |
| --- | --- |
| **Question** | **Prompts** |
| ***I would like to ask you some questions about commissioning and implementing new dementia-friendly services...*** |  |
| 9. What are the main barriers to commissioning or implementing new exercise- and activity-based interventions for people with dementia into clinical practice? | • |
| 10. What are the main facilitators for commissioning or implementing new exercise- and activity-based interventions for people with dementia into clinical practice? | • |

| **E. LONG-TERM SUSTAINABILITY** |  |
| --- | --- |
| **Question** | **Prompts** |
| ***I would like to ask you some questions about the long-term sustainability of dementia-friendly services...*** |  |
| 11. What do you think it would take to make an exercise- and activity-based intervention for people with dementia sustainable as a service? | • Who should pay for it in the future?<br>• Could it be self-funding?<br>• Is the delivery model sustainable?<br>• Is there a demand for it? |
| 12. How do you think the PrAISED training could be delivered? | • Who could provide the training?<br>• Format and structure e.g., face to face or e-training/digital?<br>• Content?<br>• Frequency/refreshers sessions?<br>• Community of Practice? |
| 13. Which professional groups do you believe will be able to deliver new exercise- and activity-based interventions for people with dementia? | • Please give examples of who e.g., exercise instructors? |
| 14. Are you aware of any dementia-friendly services that patients could be referred to after completing an exercise- and activity-based intervention to support long-term participation in activity? | • [If Yes] Can you please describe these? |

| F. CLOSING QUESTIONS |  |
| --- | --- |
| Question | Prompts |
| 15. Is there anything we haven't covered that you think we ought to know about commissioning, promoting or delivering PrAISED in routine practice or what we have discussed today? | • What advice would you give to the provider of a new exercise- and activity-based intervention for people with dementia in relation to getting the intervention commissioned, supported and promoted? |
| 16. Is there anyone else who you think we should interview in relation to this topic? |  |

## Footnotes

i One participant identified television actress Vicky McClure who had worked extensively with a dementia choir, increasing awareness of the condition (see https://www.ourdementiachoir.com/about-the-choir).

